# Measuring Integrated Novel Dimensions in Neurodevelopmental and Stress-related Mental Disorders (MIND-Set): a cross-sectional comorbidity study from an RDoC perspective

**DOI:** 10.1101/2021.06.05.21256695

**Authors:** Philip F.P. van Eijndhoven, Rose M. Collard, Janna N. Vrijsen, Dirk G.M. Geurts, Alejandro Arias-Vasquez, Arnt F.A. Schellekens, Eva van den Munckhof, Sophie C.A. Brolsma, Fleur A. Duyser, M. Annemiek Bergman, Jasper van Oort, Indira Tendolkar, Aart H. Schene

## Abstract

**Background:** It is widely acknowledged that comorbidity between psychiatric disorders is common. Shared and diverse underpinnings of psychiatric disorders cannot be systematically understood on the basis of symptom-based categories of mental disorders, which map poorly onto pathophysiological mechanisms. In the MIND-Set study, we make use of current concepts of comorbidity that transcend the current diagnostic categories. We test this approach to psychiatric problems in patients with frequently occurring psychiatric disorders and their comorbidities (excluding psychosis). The main objective of the MIND-Set project is to determine the shared and specific mechanisms of neurodevelopmental and stress-related psychiatric disorders at different observational levels.

**Methods:** This is an observational, cross-sectional study. Data from different observational levels as defined in the research domain criteria (RDoC; genetics, physiology, neuropsychology, system level neuroimaging, behavior, self-report and experimental neurocognitive paradigms) are collected over four time points. Included are adult (≥ 18 years), non-psychotic, psychiatric patients with a clinical diagnosis of a stress-related disorder (mood disorder, anxiety disorder and/or addiction disorder) and/or a neurodevelopmental disorder (ASD and/or ADHD). Individuals with no current or past psychiatric diagnosis are included as controls. Data collection started in June 2016 with the aim to include a total of 650 patients and 150 healthy controls by 2021. The data collection procedure includes online questionnaires and three subsequent sessions with 1) Standardized clinical examination, physical examination, and blood sampling; 2) Psychological constructs, neuropsychological tests, and biological marker sampling; 3) Neuroimaging measures.

**Discussion:** The MIND-Set study enables us to investigate the mechanistic underpinnings of non-psychotic psychiatric disorders transdiagnostically. We will identify both shared and disorder-specific markers at different observational levels that can be used as targets for future diagnostic and treatment approaches.

## Background

It is widely acknowledged that comorbidity between psychiatric disorders is the rule rather than the exception. Shared and diverse underpinnings of psychiatric disorders cannot be systematically understood on the basis of symptom-based categories of mental disorders, which map poorly onto pathophysiological mechanisms. In the MIND-Set study (Measuring Integrated Novel Dimensions in Neurodevelopmental and Stress-related Mental Disorders), we take advantage of concepts of comorbidity that transcend the current diagnostic categories in a naturalistic cohort of patients with frequently occurring psychiatric disorders and their comorbidities (excluding psychosis). The main objective of the MIND-Set project is to determine the shared and specific mechanisms of neurodevelopmental and stress-related psychiatric disorders at different observational levels. In the background section, we will motivate our approach generally and with respect to the choice of patients we will include.

### Current approaches in diagnosing psychiatric comorbidity

Comorbidity is not well covered by categorical, symptom-based diagnostic systems. The use of criteria to classify patients based on verbal report and observable behavior has greatly increased the reliability of psychiatric diagnoses, which serves its ultimate clinical goal of guiding treatment decisions (1, 2). However, the DSM-5’s descriptive and atheoretical approach encourages multiple diagnoses (3) and has also contributed to a conceptualization of psychiatric disorders as distinct entities that should be treated according to clinical guidelines drafted for distinct disorders. Clinical practice shows that patients with the same diagnostic classification may require different treatments, while different disorders are often treated with the same interventions, indicating that a categorial approach may overlook both heterogeneity and transdiagnostic dimensions of psychopathology. Relatedly, a large body of research indicates that factors of risk and resilience for psychopathology are not unique for distinct disorders that are identified based on symptom criteria, but commonly impact across diagnostic borders (4).

Not surprisingly in the light of the aforementioned controversy and the common dimensions, to date no biological markers have been identified that are uniquely associated with specific disorders (5, 6). Conversely, diagnostic categories seem to link poorly to underlying neurobiological mechanisms, which may better map onto dimensional diagnostic approaches that incorporate the heterogeneity of psychiatric disorders. Searching for discrete etiology underlying categorical disorders is a dead end, especially in light of the common comorbidity between disorders. Psychiatric disorders and their comorbidity should be more properly understood in a multi-dimensional, empirical framework, paving the way for new ways of understanding pathophysiological mechanisms of psychiatric disorders (7). It requires a transdiagnostic perspective that regards psychiatric disorders as related disorders with distinct and shared underlying pathophysiological pathways. As is clearly illustrated by the focus of the MIND-Set study on highly prevalent neurodevelopmental and stress-related disorders that are separable diachronically, it also requires a life span and developmental perspective that distinguishes between trait and state characteristics of psychopathology.

### Comorbidity between neurodevelopmental and stress-related disorders

In the present cohort we focus on commonly occurring comorbidities that present a challenge in diagnostics as well as treatment. Comorbidity between neurodevelopmental disorders, such as autism spectrum disorders (ASD) and attention deficit hyperactivity disorders (ADHD), and stress-related disorders, such as mood, anxiety, and substance use disorders, is common in clinical practice (8). Notably, comorbidity may also occur across the lifespan suggesting a pleiotropic genetic background of common psychiatric disorders. Comorbidity is more prevalent than would be expected by chance alone, indicating that neurodevelopmental disorders may share pathophysiological mechanisms with stress-related disorders and/or pose a risk factor for these disorders over time. Comorbidity is associated with a higher level of functional impairment and a poorer mental health outcome (9). At the clinical level, psychiatric comorbidity raises several questions related to complicated recognition and diagnosis and poses therapeutic dilemmas about the most optimal treatment strategy for particular comorbidities (10). Are depressive symptoms in someone with an autism spectrum disorder comparable to depressive symptoms in someone without an autism spectrum disorder? And at the pathophysiological level: Are these depressive symptoms related to for example biases in information processing, comparable to negative biases in major depressive disorder (MDD) without an autism spectrum disorder, which can be targeted with interventions such as Cognitive Behavioral Therapy (CBT), or should treatment for the comorbid condition be modified, and if so, how? How well is someone with an autism spectrum disorder actually able to recognize and verbalize their mood symptoms, and how does this impact the diagnostic procedure, and the treatment choice and course? And if the recognition of mood symptoms is compromised, for example when a patient shows alexithymia, how does this affect their vulnerability to stress? For ADHD, related questions arise, such as how to distinguish core attentional deficits from concentration problems related to depression? Or when do symptoms of emotional dysregulation, which are frequently observed in ADHD but not part of the formal criteria, substantiate a separate diagnosis? If so, what are the therapeutic consequences, if any? Currently, we treat comorbid depression and autism and/or ADHD mostly as solid entities that receive separate treatments, while they may share neurobiological mechanisms that may demand different targets for treatment.

### Comorbidity within the RDoC framework

High comorbidity among supposedly distinct classifications motivated the development of dimensional systems to characterize the complexity of psychiatric illness (11, 12). Trying to overcome the limitations of categorical descriptive classifications, we hence link to the Research Domain Criteria (RDoC) to study the comorbidity of neurodevelopmental and stress-related disorders (see Figure 1). RDoC offers a research framework for understanding mental disorders in terms of varying degrees of dysfunction along basic dimensions of biological systems that have been elucidated by neuroscience. Its focus on transdiagnostic mechanisms of mental disorders is rooted in a matrix with different functional domains and within domain constructs, across multiple units of analysis. Brain circuits have a central place in the units of analysis, as mental disorders are primarily regarded as disorders of the brain, which can be identified with the methods of clinical neuroscience (7). The ultimate goal of RDoC is to find biosignatures that on the one hand improve current diagnostic approaches (13), and on the other hand help to understand the working mechanisms of existing therapeutics and serve as targets for new treatments.

**Figure 1.**
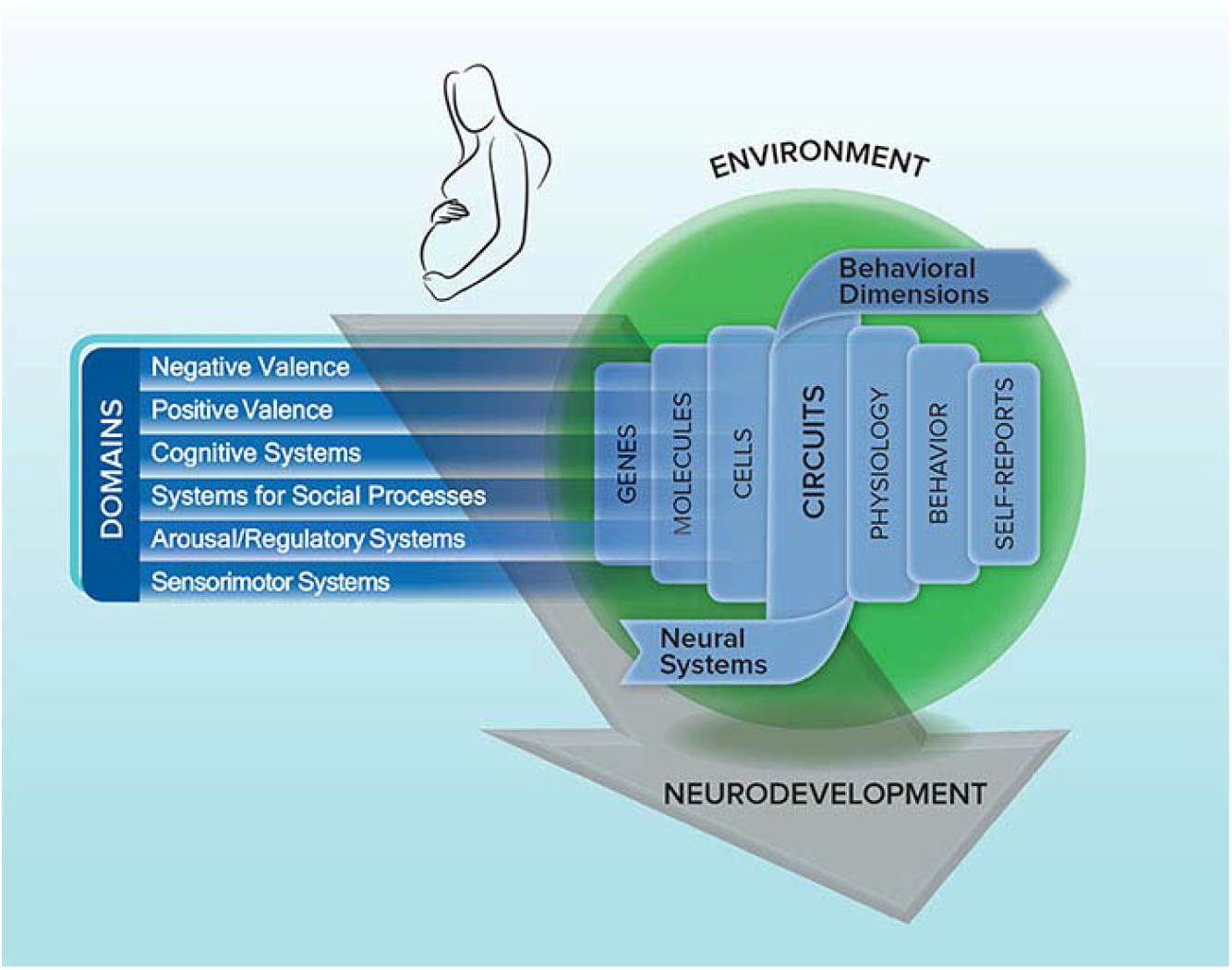
An overview of the RDoC framework (https://www.nimh.nih.gov) ^13^

Six functional systems are identified that serve the basic motivational and adaptive needs of an organism: The negative and positive valence systems, cognitive systems, arousal and regulatory systems, social processes, and sensorimotor systems. The negative valence system directs responses to aversive stimuli or contexts, whereas the positive valence system addresses such responses to positive situations. The cognitive system contains various cognitive processes such as memory and cognitive control, whereas social processes mediate the responses to interpersonal settings. Arousal and regulatory systems include processes that are responsible for the activation of neural systems within certain contexts, as well as homeostatic regulation. Sensorimotor systems are involved in motor behaviors. Each domain contains up to seven constructs, such as “acute threat” and “loss” in the negative valence system and “affiliation and attachment” and “perception and understanding of self” in the social processes system. These constructs and domains are to be analyzed with different methods and at different units of analysis: from a genetic, molecular or cellular level to neural or brain circuitry and further to the physiological and behavioral level, onwards to the level of self-report and paradigms.

The RDoC framework fits ideally with our purpose to increase understanding of psychiatric comorbidity by studying neurodevelopmental and stress-related disorders in terms of distinct or related underlying vulnerability factors and etiopathogenetic mechanisms. By considering the multiple levels of analysis, the RDoC inspired MIND-Set study aims to integrate various biological and behavioral measures and analyze constructs and mechanisms dimensionally, across a spectrum of functioning that is not restricted by the boundaries of distinct psychiatric disorders. Last but not least, the RDoC approach stresses the importance of the neurodevelopmental and environmental context of behavior and biological systems, which is an essential aspect for studying comorbidity within the current project.

Several RDoC domains seem particularly relevant for advancing our understanding of the underlying mechanisms for comorbidity of neurodevelopmental and stress-related disorders: cognitive systems, negative valence, positive valence, social processes and arousal and regulatory systems. Below we specifically focus on the importance of cognitive systems as well as negative and positive valence.

*Cognitive systems* include perception, attention, cognitive control and (working) memory. The cognitive systems domain is rarely studied in isolation; its specific constructs are considered as underlying to cross-domain processes related to psychiatric disorders. Both neurodevelopmental and stress-related disorders have important cognitive characteristics, such as deficits in executive functioning, emotion processing and emotion regulation, which have been recognized to determine psychopathological symptoms and behavior. Recent neurocognitive findings suggest that problems in emotion regulation result from preferential processing of (negative) emotional information in subcortical structures including the amygdala, and reduced prefrontal executive control to inhibit inappropriate emotions and emotion expression (e.g. (14, 15)). Indeed, increased and biased processing of emotional information has been demonstrated extensively in stress-related disorders (16-19) and contributes to the onset, maintenance and relapse of these disorders (20-24). In neurodevelopmental disorders, emotional information processing has been studied less frequently and findings are conflicting with regard to the presence of potential negative biases (25-29).

In our mechanistic approach across a range of neurodevelopmental and stress-related disorders, both executive functioning (cognitive systems) and emotional information processing (negative valence systems) are key mechanistic elements, which may interact in specific ways. The function of these covert cognitive mechanisms in several cross-disorder symptoms such as impulsivity, apathy or alexithymia are yet unknown. Characterizing these mechanisms may allow us to identify different underlying profiles that combine executive dysfunction and emotional processes biases and could serve as targets for new treatments such as neuromodulation. A specific example, that may illustrate partly overlapping mechanisms, is a deficit in mental shifting that may be implied in preoccupied and rigid thinking that is characteristic for ASD, but which is also implied in the ruminative thinking that characterizes depression. In individuals with ASD there is some evidence that poorer executive functioning (and greater behavioral inflexibility) predicts greater anxiety and depression (30, 31). Similarly, executive deficits have been related to rumination (32) and the susceptibility to depression (14). Relatedly, ADHD and depression share related aspects of executive dysfunction such as attentional deficits and difficulties in decision making (33).

*The negative valence systems* are responsible for the responses to aversive situations or context, and lead to the experience of negative affect, such as sadness, anxiety and anger. As such, negative emotions are the hallmark of stress-related disorders such as depression and anxiety disorders, characterized by persistent patterns or high levels of negative affect. The subjective domain of negative affect is accompanied by objective biases in information processing in the form of attentional and memory biases. Negative affect, such as depressed mood and anxiety, both on a symptomatic and syndromic level, is frequently comorbid in neurodevelopmental disorders. ASD is a neurodevelopmental disorder primarily characterized by alterations in sensory sensitivity, inflexible routines, restricted interests and deficits in social functioning, but about 50% of high-functioning adults with ASD who were referred to a psychiatry department had comorbid MDD (34). One possibility is that negative affect in ASD results from increased levels of stress sensitivity that are related to these primary deficits, for example increased levels of stress caused by sensory overstimulation or problems in relationships related to deficits in social cognition and flexibility (35). Individuals with ASD and ADHD may also be more vulnerable to depression and anxiety because they share information processing styles that are related to the susceptibility for depression and anxiety, such as biases in information processing (36).

With regard to comorbidity, we know that later in life, individuals with ASD have a four times higher lifetime prevalence of depression. Because of the overlap of symptoms and personality characteristics (e.g. rigidity) depression is often difficult to recognize in ASD and remains frequently undetected (37). Individuals with ASD have difficulties reading their own inner states and clinicians lack diagnostic tools and treatment options. Recognition and treatment is needed as individuals with MDD and ASD have lower global functioning compared to individuals with ASD only. Our understanding of MDD in ASD remains however limited today, as well as our treatment options. One leading theory for the development and maintenance of depression is Beck’s idea of dysfunctional assumptions (resulting from negative life experiences) that organize into cognitive schemata, and when activated by stress result in cognitive biases (38). These biases have been found in a range of domains (e.g. attention, perception, interpretation, and memory (19, 39)) and are not uniquely related to depression; they are also detected in anxiety and addiction disorders (40-43), and have even been related to early onset neurodevelopmental disorder symptoms such as ADHD (29).

*The positive valence system* addresses the systems underlying the ability to experience feelings of pleasure. Anhedonia as a hallmark of depression is more likely in persons with a reduced ability to experience pleasure (low hedonic tone), which is associated with ADHD and substance abuse, explaining patterns of comorbidity that can be tracked down to commonalities in the underlying neurocircuitry that regulates emotional affect and reward processing or even define subtypes of depression that are relevant for treatment outcome (44). Moreover, in the context of our interest in assessing negative biases, it is also important to understand whether participants with depression will exhibit enhanced learning in the face of punishment in comparison to reward (45).

### Data-driven approaches

In the light of the different levels of data acquired within the RDoC framework, we can disentangle the concept of psychiatric comorbidity by data-driven approaches that are not constrained by the clinical categories. Moreover, as working principally from the RDoC perspective means working back and forth through different domains and analysis units (e.g. linked independent component analysis), we will aim to find cross-domain links with still data-driven procedures, with only at the end of the endeavor assessing the relation to clinical categories, including the descriptive comorbidities.

MIND-Set, our innovative cross-sectional cohort study, has to be understood as a further step in understanding comorbidity from an RDoC perspective by including patients classified with neurodevelopmental disorders with an early age of onset (ASD: 1-5 years, ADHD: 5-12 years) and/or stress-related disorders with on average an adult age of onset. We include patients with at least one of these broadly used classifications, aiming to study underlying shared and distinct mechanisms. By allowing for both the DSM-5-based classifications of disorders and new to-be-developed ordering of symptoms and behavior, we push the psychiatric research field forward while also fostering a close translation to clinical practice.

MIND-Set does not involve longitudinal changes directly (e.g. improvement of prognosis through interventions) in our patients, which is the step to be taken to leverage these insights to clinical practice and which will be addressed by planned follow-up studies. The advanced understanding of comorbidity resulting from this initial study will be an essential stepping stone in order to progress towards innovative ideas about new therapeutic approaches that in the end will hopefully change clinical practice for patients suffering from a multiplicity of symptoms. Nonetheless, we include an online survey including questionnaires on anxiety, mood, stress adaptation, and daily functioning to monitor psychiatric variables during the COVID-19 pandemic in our cohort.

### Study aims and outline

The main **objective** of the MIND-Set study is to determine the shared and specific mechanisms of neurodevelopmental and stress-related psychiatric disorders at different observational levels to gain insight in the comorbidity of the most common non-psychotic disorders (i.e. neurodevelopmental and stress-related disorders).

We will realize this aim by adopting a dimensional approach focusing on dysfunction related to stress-related (mood, anxiety and substance abuse) and neurodevelopmental (autism, ADHD) disorders. This will allow us to investigate connections between different units of analysis (connect symptoms with underlying circuits) and derive profiles that improve current understanding of comorbidity and ultimately can lead to better treatment.

## METHODS

### Design

The MIND-Set study is an observational, cross-sectional study, in which data from different observational levels according to the research domain criteria (RDoC) units of analysis (genetics, physiology, neuropsychology, system level neuroimaging, behavior, self-report and experimental neurocognitive paradigms) are collected over four time points for patients with neurodevelopmental and stress-related disorders and healthy controls.

### Setting

The MIND-Set study is mainly executed at the outpatient unit of the psychiatric department of the Radboud university medical center (Radboudumc), Nijmegen, the Netherlands. The department specializes in the diagnosis and treatment of neurodevelopmental disorders and stress-related disorders in adults, with a special attention and expertise for psychiatric comorbidity and combined psychiatric and somatic pathology. Inpatients who are able to be investigated can also participate in the study.

### Population

#### Patients

##### Inclusion criteria

Included are adult (≥ 18 years) psychiatric patients with a clinical diagnosis of a stress-related disorder (mood disorder, anxiety disorder and/or addiction disorder) and/or a neurodevelopmental disorder (ASD and/or ADHD).

##### Exclusion criteria

Patients with diseases of the central nervous system resulting in (permanent) sensorimotor and/or (neuro)cognitive impairments, a current psychosis, a full scale IQ estimate < 70, inadequate command of the Dutch language or who are mentally incompetent to give informed consent are excluded from participation. Additional exclusion criteria for the MRI session are: metal objects in the body (excluding dental fillings), ferromagnetic implants or pacemakers, jewelry or piercings that cannot be removed, brain surgery, epilepsy, claustrophobia, pregnancy and self-declared inability to lie still for more than one hour.

##### Control participants

Individuals with no current or past psychiatric diagnosis are included. Possible eligible individuals are approached via databases of the department’s previous studies, advertisement in newspapers, social media, websites, and via the research participation system of the Radboud University Faculty of Social Sciences (‘SonaSystem’), as well as verbally through the researchers’ own networks. The absence of lifetime psychiatric diagnoses is assessed via a telephone screening interview, using the same diagnostic measurement instruments as described below for the patient sample.

##### Numbers

Data collection started in June 2016. We aim to include a total of 650 patients and 150 healthy control participants in 2022.

### Procedure

The data collection procedure includes an online assessment and three subsequent sessions:

- Online assessment: online self-report questionnaires assessing demographics, symptomatology and functioning
- Session 1: Standardized clinical examination, physical examination and blood sample
- Session 2: Psychological constructs, behavioral tasks and neuropsychological tests, biological markers
- Session 3: Neuroimaging measures

The procedure for each part is briefly described below. An overview is given in Table 1, including the full names of the measurement instruments used. In the last column of Table 1, we categorize the data according to the six units of analyses as proposed by RDoC (self-report, behavior, physiology, circuits, cells and molecules).

**Table 1:**
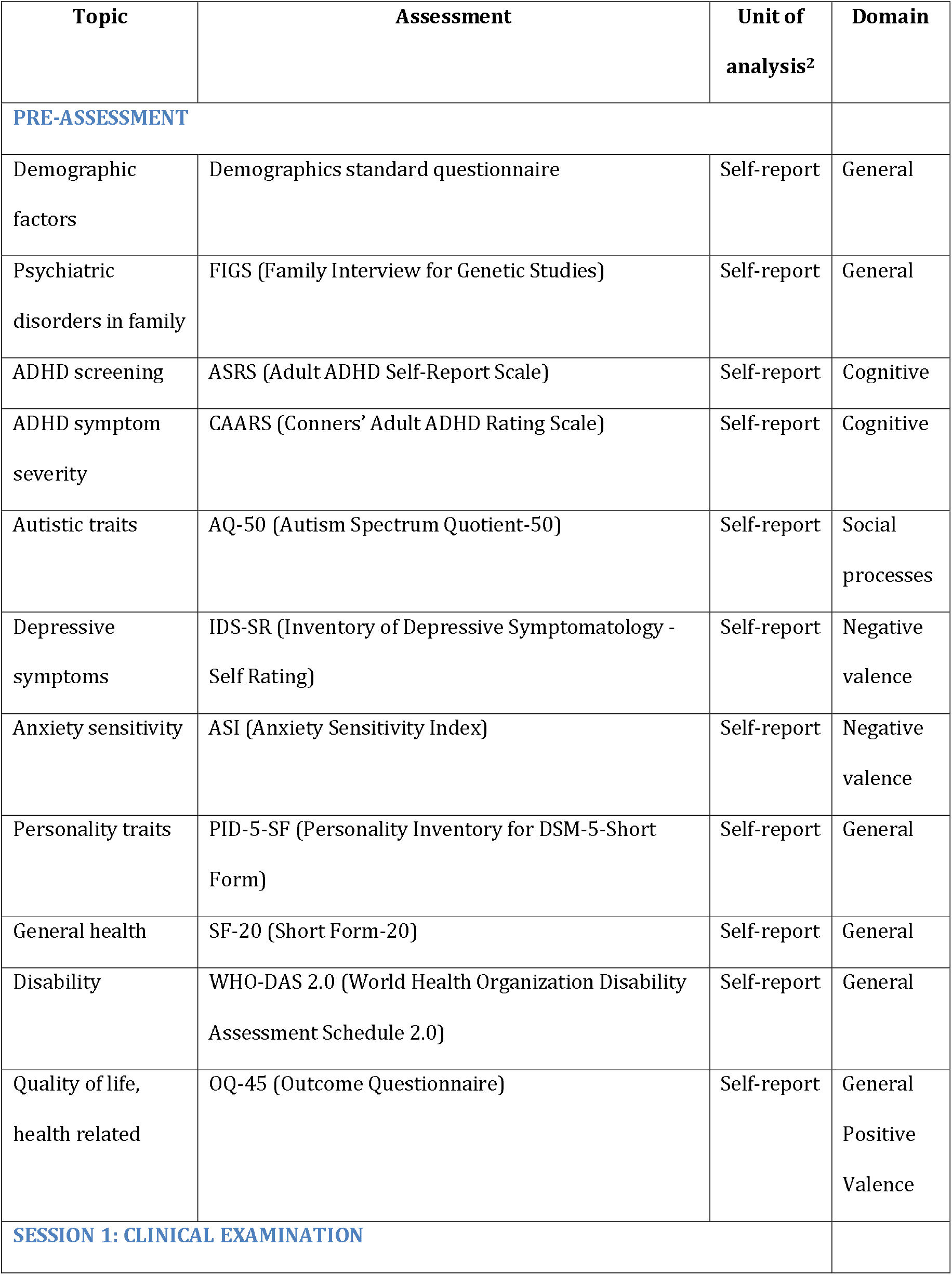
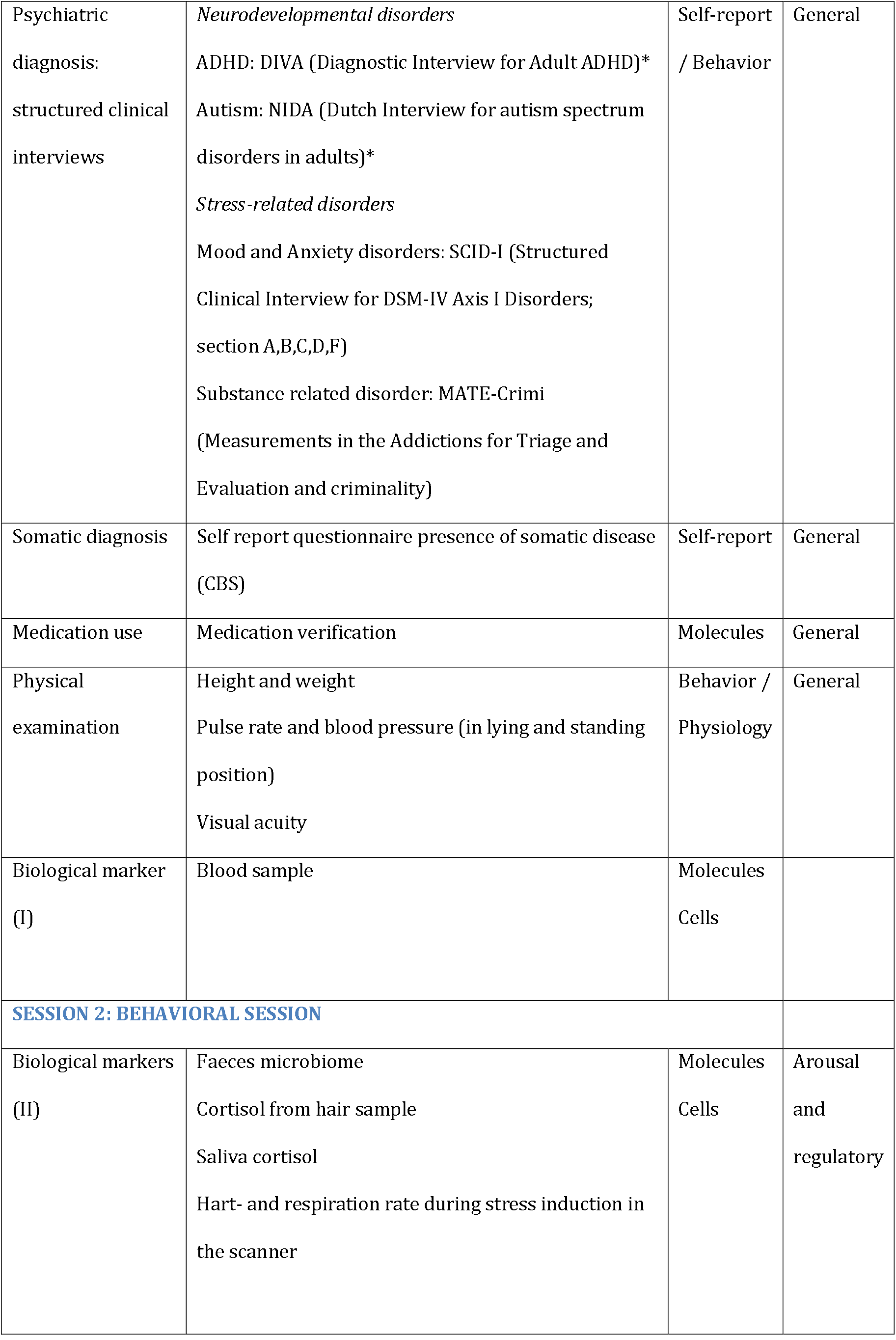
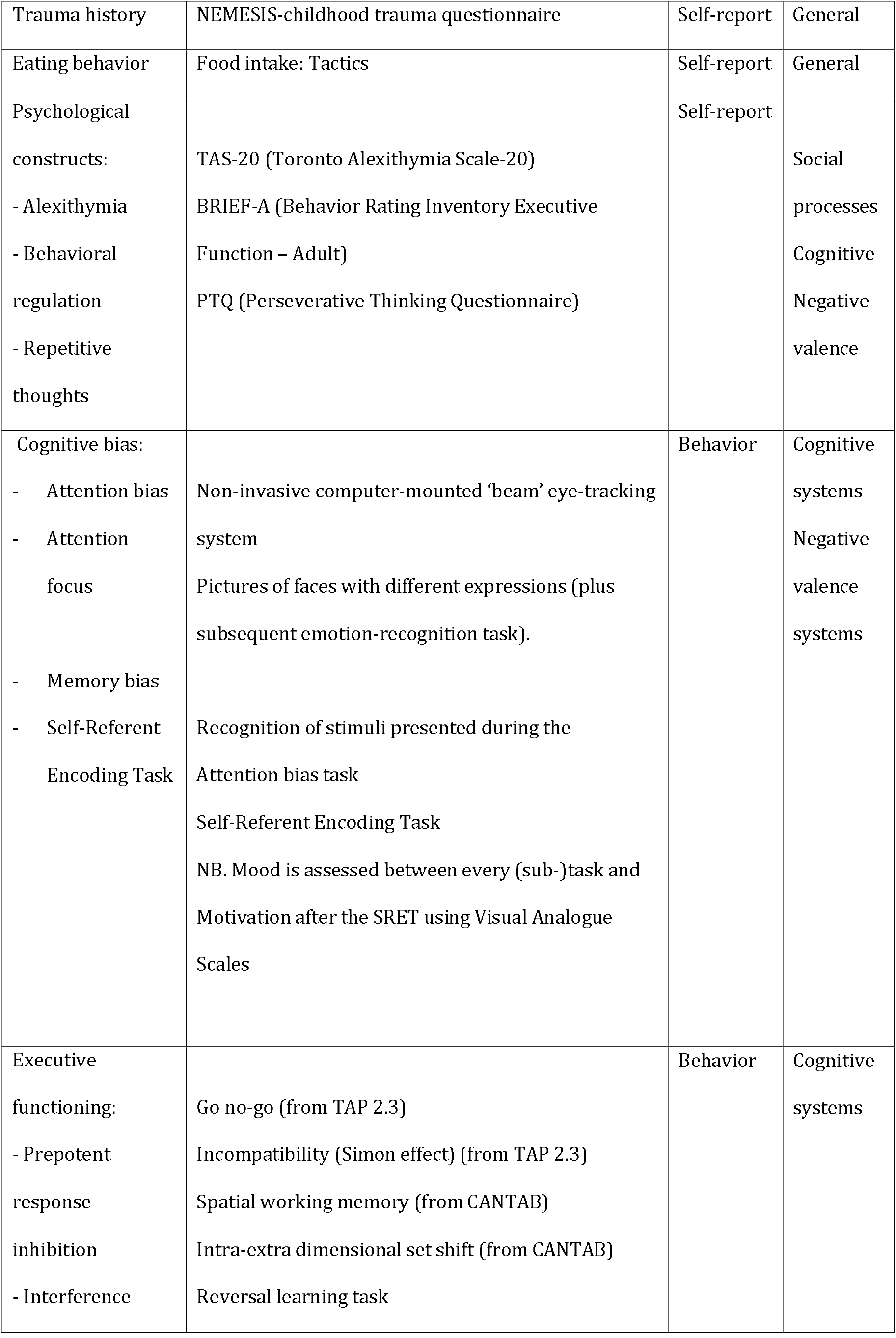
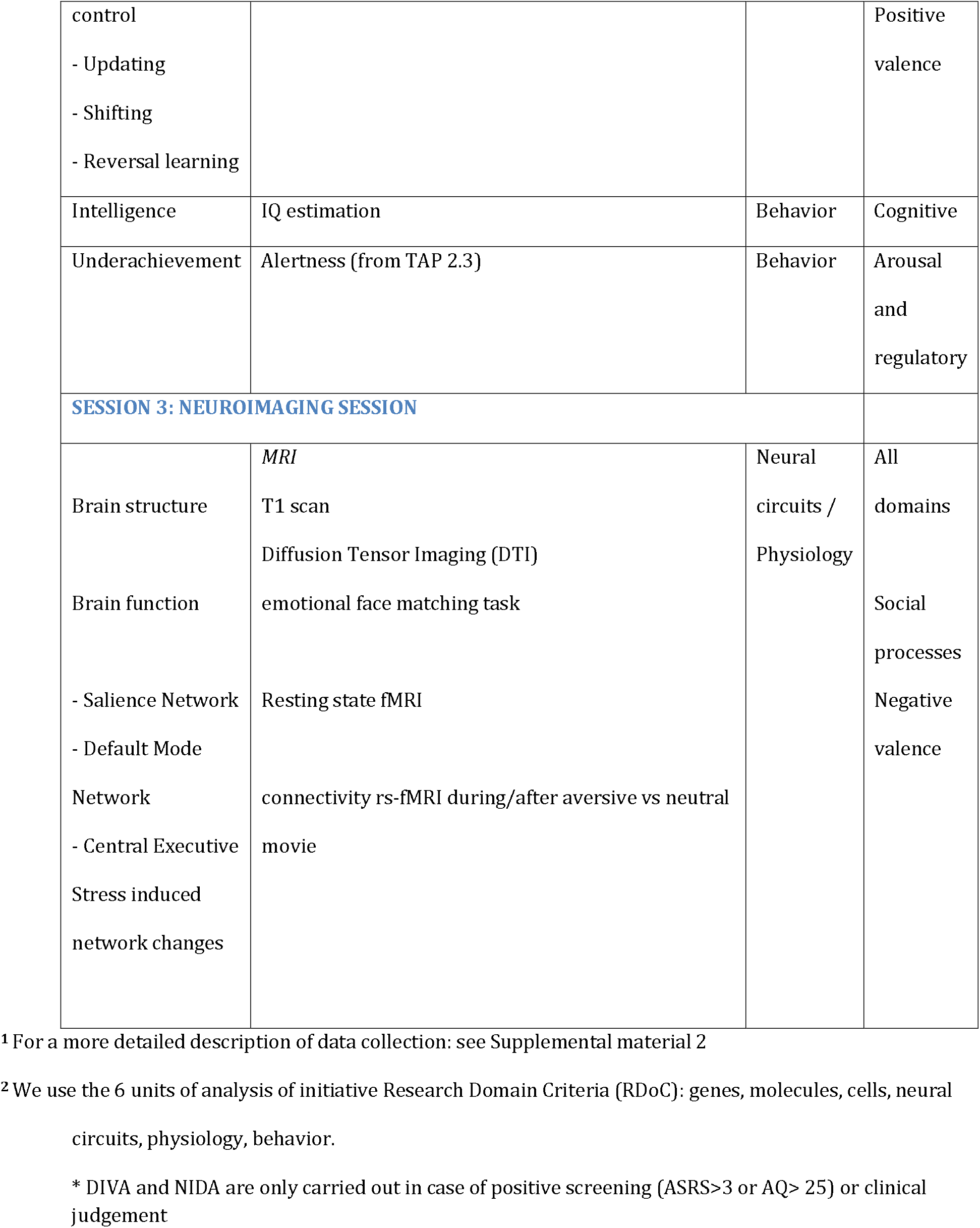
Data collection of the MIND-Set study: topics and instruments ^1^

### ONLINE ASSESSMENT

#### Questionnaires

all patients referred to the outpatient psychiatric department receive login details for an online questionnaire batch at home. They are asked to fill out the questionnaires within 21 days before their first appointment. If preferred, a paper copy is sent to their home address. The questionnaires assess demographics, psychiatric disorders in the family, symptoms of ADHD, depression and anxiety, and autistic and personality traits. Two questionnaires are also used as screening instruments for autism and ADHD. Finally, questionnaires on general health, disability or functional limitations and quality of life are included. Summary and subscale scores derived from these questionnaires are made available before the clinical examination session to inform the clinician about the possible involvement of neurodevelopmental and stress-related disorders, personality problems and functional status.

## SESSION 1: CLINICAL EXAMINATION

### Diagnostics

during a three-hour clinical examination at the psychiatric department, patients undergo a psychiatric, biographical and somatic anamnesis, medication verification, review of treatment history, structured clinical interviews, a physical examination and a questionnaire assessment of the presence of somatic diseases. Examinations are conducted by well-trained clinicians: psychiatrists, psychologists, supervised psychiatric residents, supervised nurse practitioners and supervised psychology interns. At the end of the examination, the senior clinician assesses eligibility based on the DSM classification (see below) and completes the written informed consent procedure. The patient consents to a) the use of their questionnaire data for research, b) the use of their diagnostic data for research and c) participation in the next sessions of the study. After giving informed consent, blood sampling is executed and appointments for Session 2 and 3 are scheduled to take place as soon as possible and ultimately within 90 days

## SESSION 2: BEHAVIORAL SESSION

### Biomarkers

first, patients receive a package and instructions for the collection of a faeces sample at home. They are instructed on how to return this package by mail. Next, hair samples are taken for cortisol (and possibly other hormones) measurement.

### Questionnaires and neuropsychology

first, patients undergo a neuropsychological assessment (∼120 minutes), including a pen and paper task and several computer tasks including an eye-tracking task. The test battery is administered by a trained research assistant. Then, subjects are required to fill out questionnaires (∼20 min) assessing trauma history, food intake and three psychological constructs (alexithymia, repetitive thoughts and behavioral regulation). A research assistant is available for assistance.

## SESSION 3: NEUROIMAGING SESSION

This final session (180 min) is scheduled in the afternoon to account for the diurnal changes in cortisol levels at the Centre for Cognitive Neuroimaging of the Donders Institute for Brain, Cognition and Behavior in Nijmegen. It starts with an acclimatization period during which participants fill in questionnaires about current mood state, recent medication changes and watch a relaxing nature documentary. Hereafter, they are prepared for the MRI scanner and undergo different imaging paradigms including a T1 structural MRI, DTI, functional MRI during an emotion-recognition task and a baseline resting state fMRI. It continues with resting state fMRI after a neutral and a highly aversive movie clip, meant as a brief stress induction procedure. During the whole imaging session physiological data are collected such as heart and respiration rate, and saliva for cortisol and alpha-amylase measurement is collected at different time-points in addition to assessments of mood, stress-level and other emotions. The neuroimaging session ends with a short debriefing procedure.

### Measures: some specification

After the description of the logistics of our data collection, we will give background information about some specific parts of it. The supplementary information offers a complete description of the specific instruments and measures.

#### Psychopathology

Psychopathology is addressed along a continuum ranging from the syndrome or disorder level (DSM-IV and DSM-5) to the disorder-related symptomatic level, and to the transdiagnostic dimensional level. This in order to provide information about the primary diagnoses, and the core symptom dimensions and psychological constructs that accompany these diagnoses.

#### Disorders

For each of the different neurodevelopmental and stress-related disorders, state-of-the-art diagnostic instruments were selected to classify DSM-IV or DSM-5 diagnosis^1^. Neurodevelopmental disorders are assessed in case of either positive screening or based on clinical judgment by diagnostic interviews. For screening on ASD traits we use the Autism Spectrum Quotient (AQ-50) (46). When a patient scores positive on this instrument (50 items, cut-off >25) we next use the Dutch Interview for the Diagnosis of ASD in adults (NIDA) (47) to diagnose ASD according to the DSM-5. Regarding ADHD, we use the World Health Organization Adult ADHD Self-report Scale (ASRS)-short version for screening (48). In case of positive screening (6 items, cut-off >3), we subsequently conduct the Diagnostic Interview for ADHD in adults (DIVA) (49) to diagnose ADHD according to DSM-IV. Both the DIVA and NIDA are completed in the presence of a partner and/or family member of the patient (if available) to be able to retrospectively and collaterally ascertain information on a broad range of symptoms in childhood and adulthood.

Stress-related disorders are standardly assessed with the Structured Clinical Interview for DSM-IV Axis I Disorders (SCID-I) and the Measurements in the Addictions for Triage and Evaluation and criminality (MATE). The SCID-I (50) is used to diagnose mood (depression and anxiety) disorders and to exclude psychotic disorders. To diagnose substance related disorders according to DSM-5, we use an adapted version of the MATE (51).

#### Symptoms

A set of questionnaires provide measures of depression (IDS), anxiety (ASI) and ADHD symptoms (CAARS) to provide dimensional measures that fit with the syndromes that are our primary diagnoses, but also to assess comorbidity at the symptomatic level in the context of other diagnostic categories.

#### Traits

We use the Personality Inventory for DSM-5 (PID-5-BF) to assess personality trait domains including negative affect, detachment, antagonism, disinhibition, and psychoticism and the AQ-50 to measure traits that are related to autism in adults with normal intelligence.

#### Psychological constructs

We have included three questionnaires that address psychological constructs that cut across syndromes and reveal transdiagnostic mechanisms important for understanding comorbidity. We include the perseverative thinking questionnaire (PTQ) and alexithymia (TAS-20) and behavioral regulation (BRIEF-A) questionnaires. In addition, a structured inventory developed for the NEMESIS epidemiological study assesses an individual’s trauma history before the age of 16 including emotional neglect, psychological, physical and sexual abuse (52, 53).

#### Somatic status

Data on somatic health are derived from physical examination, medication review, somatic anamnesis and questionnaires about somatic status and functional limitations. A physical examination including height, weight, pulse rate and blood pressure measurements (the latter in both lying and standing position) is conducted during the first clinical appointment. Current medication use is determined by medication verification using both an up-to-date list of medication from the pharmacy and information from the patient about current use. A somatic anamnesis of diseases is assessed by using the Statistics Netherlands questionnaire (CBS) (54) that informs about the presence of a wide range of somatic diseases. General health is assessed by the short form of the general health survey (SF-20), which is a multidimensional instrument assessing different domains of health. Functional limitations are assessed with the self-report WHO-Disability Assessment Schedule 2.0 (WHODAS) (55), covering six different domains of functioning. The self-report Outcome Questionnaire-45 (OQ-45) measures distress associated with mental health problems across disorders and assesses functioning in interpersonal relations and social roles (56).

#### Biomarkers

#### Blood sampling

Blood samples are taken from each patient during the clinical appointment. Assays include measures of hematology, electrolytes and endocrine function. We also collect blood for the analysis of inflammatory markers, immune function and for further studies in genetics (DNA, RNA) and epigenetics as outlined below:

#### Peripheral blood protein biomarkers

In peripheral blood, a large panel of proteins, representing neurotrophic, immunological and metabolic markers, will be profiled by the implementation of quantitative immunoassays or proteomics, using multiplex enzyme-linked immunosorbent assay (ELISA) approaches. For this analysis, we will use multiplex ELISA panels including chemokines, cytokines, inflammation markers, growth factors and/or metabolic markers.

#### DNA polymorphisms and genotyping

Genome-wide profiling of single nucleotide polymorphisms, copy number variants, insertions and deletions will be performed using microarray platforms. Genotyping will be conducted on DNA isolated from peripheral blood mononuclear cell (PBMCs).

#### DNA methylation

Genome-wide profiling of differential DNA methylation status will be analysed using the Illumina Infinium MethylationEPIC beadchip microarray platform in DNA isolated from PBMCs. Standardized protocols such as MethylAids or RnBeads, will be used for the processing of raw data and downstream data analyses. The pipeline choice will be made prior to data analysis, based on the current state of the art.

#### Scalp hair

Scalp hair is used for cortisol measurement. Hair cortisol provides an index measure of HPA axis activity over the last 3 months (57), much like HBA1c is used as a proxy for the mean level of glucose in the past 3 months. As such, the hair cortisol gives a more persistent marker of the levels of chronic stress associated with negative emotions.

#### Saliva

Saliva is collected at six time-points during the imaging session (baseline, right before scanning, after the structural T1 scan, before and after stress induction and when scanning is finished) and twice at home for a total of eight samples to probe a cortisol curve both in a relatively stress free situation and under conditions of stress.

#### Microbiome

The relationship between the bacterial communities in the gut (the microbiome) and the brain is bi-directional: the brain alters the gut microbiome, and the gut microbiome also modulates brain function. Studies conducted over the last decade in rodents show that gut microbiota can modulate brain development, neurotransmitter systems, signaling pathways, and synaptic proteins, converging in observable behavioral changes. Recent research has pointed to a possible link between autism spectrum disorder (ASD) and the microbiome and it is well known that many patients with autism suffer from gastrointestinal symptoms (58). Recent findings also suggest that dysregulation of the microbiome is involved in the pathophysiology of stress-related disorders in general, and depression in particular (59).

#### Neuropsychological assessment

The RDoC unit behavior is operationalized by neuropsychological assessments within the domains of the negative valence systems (constructs: sustained threat, loss), positive valence systems (construct: reward learning) and cognitive systems (constructs: attention, declarative memory, cognitive control). Affective neuropsychological tests assess emotional processing and in the context of the negative valence system we focus on several cognitive biases. Here, we assess attentional bias for both social and non-social negative and positive pictures by means of a free-viewing eye-tracker task (with a non-invasive computer-mounted ‘beam’ eye-tracking system) and a subsequent recognition task to assess memory bias during eye-tracking. Measuring eye movements during a task using an eye-tracker is regarded as a reliable measure for attentional focus (60). Patients with depression show more attention towards negative information, which probably points to a difficulty to disengage from negative information (17), but in comparison with healthy individuals they also show less attention to positive stimuli (61). As patients with autism generally show decreased attention to social information (62), we have chosen to incorporate both social and non-social pictures with either negative or positive valence, in order to be able to dissociate the differential contribution of these factors on attentional processes. Secondly, memory bias is tested by a computerized self-referent encoding task (63) in which participants have to indicate how characteristic different positive and negative adjectives are to them and are subsequently tested for correct recall of these adjectives after a distraction task. Biases in information processing have traditionally been studied within the boundaries of diagnostic categories and have mainly been studied in affective disorders. Negative memory bias seems to be associated with a higher level of comorbidity among psychiatric disorders (64). Biased information processing may therefore constitute a transdiagnostic mechanism for psychopathological symptoms, which seems crucial for understanding comorbidity. As biased information processing constitutes a cognitive vulnerability that, according to Beck’s model (38, 65), is linked to the experience of adverse events during childhood, which may lead to dysfunctional cognitive schemas, we have included a structured inventory of childhood adversity (NEMESIS questionnaire)(28). This allows us to study the relationship between the subjective report of childhood adversity and objective measures of cognitive biases.

Several tests are included to measure different aspects of executive functioning, as well as a test to estimate premorbid intelligence for matching purposes. Visual analogue scales are used to assess mood at four different timepoints throughout the assessment to account for the influence of mood on performance, as well as self-reported effort on the tests afterwards.

Finally, within the domain positive valence systems, we measure the construct of reward learning. Learning can be influenced by the valence of the feedback given on the performance during the task. For example, previous studies have found reduced learning from reward in mood disorders (66-69). Here, we employ a probabilistic reversal learning task (70-72) to examine reward and punishment sensitivity in a changing context. First, participants learn a stimulus-response relationship by trial-and-error, after which the stimulus response relationship is reversed without explicit warning and they have to change their response. Reversal learning is an important aspect of cognitive flexibility, which supports someone to adapt to changing environmental conditions including rewards (73). Deficits in reversal learning have been shown in stress-related disorders (74) and evidence also points to impairments in reinforcement learning in ASD (75) which may predispose to specific patterns of comorbidity. Disrupted reinforcement learning in ADHD is probably caused by altered dopamine signaling, which is hypothesized to underlie comorbid addictive behavior (76). Performance on the probabilistic reversal learning task is dependent on both serotonergic (punishment sensitivity) and dopaminergic (reward sensitivity) effect, which can be dissociated by specific genetic polymorphisms of the respective transporter systems (72). The neural correlates of this task are well characterized by neuroimaging studies which demonstrate the recruitment of fronto-striatal circuitry during reversal learning (77).

#### Cognitive systems

Cognitive impairment and emotional regulation deficits are common in both stress-related and neurodevelopment disorders. Our aim here is to study the nature of these alterations in executive functioning by studying prepotent response inhibition, interference control, updating and shifting across stress-related and neurodevelopmental disorders, in order to better understand the underlying mechanisms of shared symptoms such as impaired emotion regulation, rigidity and impulsivity. ASD and ADHD are both associated with impairments in executive function and each disorder is thought to have its specific deficits with impairment in shifting most prominent in ASD (78) while ADHD is typically characterized by problems with behavioral inhibition (79). Evidence suggests that the level executive function is an important predictor of comorbid anxiety and depression, and that specific deficits of ASD and ADHD may reveal pathways to comorbidities in these disorders (31).

In ASD it has been shown by several meta-analyses that overall executive function is consistently impaired showing a relatively stable pattern over time, but evidence could not point to a specific profile of executive dysfunction such as shifting associated with ASD (80). Performance of executive function in ASD is thought to be related to poor regional coordination and integration of prefrontal executive processes that integrate with emotion and social circuits, reflected by aberrant patterns of connectivity with both changes of within-and between-network functional connectivity scale networks (81). A recent data-driven approach identified three transdiagnostic subtypes of executive functioning in a large sample of children with ASD, ADHD and neurotypical children, that spanned the normal to impaired spectrum but also cut across ADHD and ASD samples. Moreover, these subtypes of executive functioning better accounted for variance in the neuroimaging data than DSM diagnoses did, highlighting the point that transdiagnostic subtypes may indeed refine current diagnostic classifications (82).

#### Neuroimaging: Brain circuits

The brain circuits level is at the core of our research design, as it bears on the hypothesis that the phenotypic, behavioral differences among psychiatric disorders can be explained by differences in the underlying neural circuitry, while downstream causal mechanisms such as genetic and epigenetic effects or environmental factors will lead to psychiatric symptoms and disorders via their disruptive effects on neural circuits. The brain is dynamically organized into functional networks of interconnected areas, which interact to perform unique brain functions. These networks can be consistently identified with functional MRI scans during the “resting-state”, by calculating functional connectivity between voxels. The most relevant networks with regard to psychiatric disorders are the default mode network (DMN), the salience network (SNand the central executive network (CEN) (see figure 2).

**Fig. 2.**
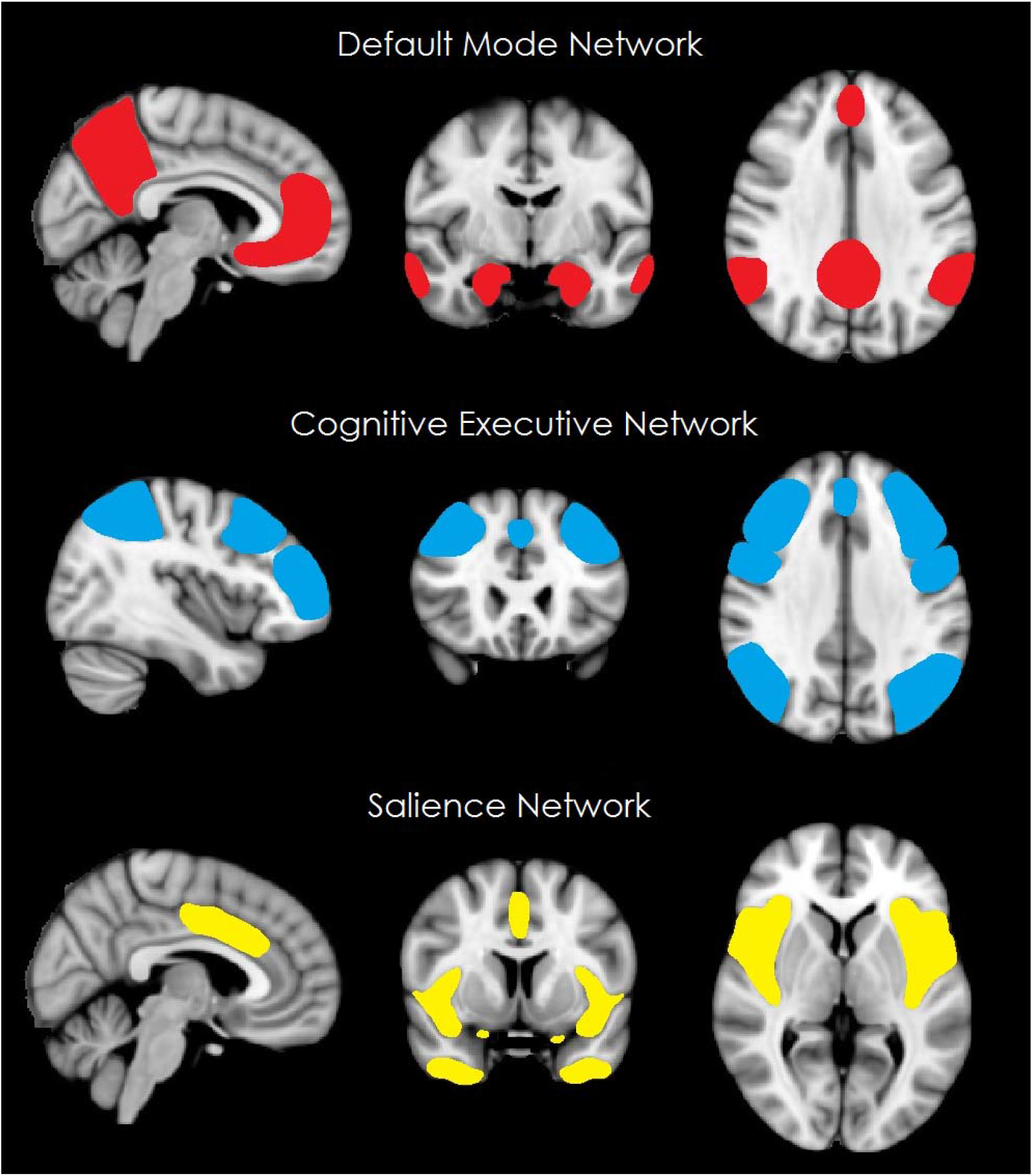
Representation of relevant resting-state networks with the default mode network depicted in red, the central executive network in blue and the salience network in yellow (adapted from^143^).

The DMN, covering the medial prefrontal cortex, anterior cingulate cortex (ACC) and precuneus (83), is most active during rest and decreases activity during demanding cognitive tasks. The DMN is related to emotion regulation, self-reference and obsessive ruminations (84). The SN, covering fronto-insular cortex, dorsal ACC and temporal poles, plays a central role in emotional control (85). Inherently linked to the SN is the affective network (AN), covering the ventral ACC, amygdala, nucleus accumbens and orbitofrontal cortex, which is involved in emotion regulation and monitoring the salience of motivational stimuli (86-88). The CEN, including lateral parts of prefrontal and parietal cortex, is most active during cognitive tasks and is relevant for attention and working memory. These networks are stable across time, but are also dynamically changing and interacting which each other, leading to different network-configurations or “connectivity states” in which these networks have specific relative contributions. Global states such as sleep or acute stress are characterized by large changes in the network-configurations. Together these networks cover the most important functional domains **such as** top down cognitive control, conflict signaling, salience detection, self referential processing that are affected in both stress-related and neurodevelopmental disorders. Small pilot studies with this approach have already demonstrated that hyperconnectivity in components of the DMN is associated with depressive symptoms such as ruminations and self-absorption while hypoconnectivity in components of the DMN is associated with anxiety symptoms (89). Studying the dynamics of network connectivity, in conditions of both rest and stress, allows us to disentangle fundamental pathophysiological mechanisms underlying these disorders and their shared mechanisms that are relevant for understanding comorbidity.

Some of the network changes may be a consequence of structural changes in white matter fiber systems. Magnetic resonance diffusion tensor imaging (DTI) evaluates both the orientation and the diffusion characteristics of white matter tracts in vivo (90) and tractography methods allow to extract white matter fiber bundles relevant for psychiatric disorders. Changes in structural connectivity provide extra parameters in the attempt to identify connectivity signatures that may reflect fundamental pathophysiological features (91). Investigations with DTI and functional MRI in parallel are still at its start and our hypotheses are preliminary at this stage. Based on anatomy we expect that diffusion measures within the cingulum fasciculus will be most relevant for connectivity patterns of the DMN, while measures of the uncinate fasciculus and fornix will be mostly associated with the SN and CEN.

##### Negative valence system

We will investigate the functional networks both during resting state and during a brief stress induction procedure (acute threat paradigm). Previous research has shown that acute stress shifts the brain into a state that fosters rapid defense mechanisms (89). Stress-related neuromodulators are thought to trigger this change by altering properties of large-scale neural populations throughout the brain. In healthy subjects, we have shown that noradrenergic activation during acute stress results in prolonged coupling within a distributed network that integrates information exchange between regions involved in autonomic-neuroendocrine control and vigilant attentional reorienting. It remains unclear to what extent these mechanisms are altered by psychiatric diseases, thereby reflecting an acute measurement of vulnerability and disease load. Functional measures will be complemented by diffusion weighted imaging to provide measures of structural connectivity between the networks. Further, we want to explore if dynamic functional connectivity data along the baseline-stress-recovery axis for the three distinct networks, will serve to identify differences in the dynamic balance in these networks at individual subject level, and can be related to behavioral and symptom profiles.

##### Social processes

An emotional face matching task (EFMT) addresses the subconstruct reception of facial communication within the social processes domain. This paradigm engages the amygdala and an amygdala-centered network by contrasting the BOLD response during blocks of angry and fearful face stimuli with blocks with geometric shapes that consist of scrambles of the same face stimuli (92, 93). This task is commonly used as a paradigm to probe amygdala reactivity and aberrant amygdala reactivity has been implicated in both stress-related and neurodevelopmental disorders. Neuropathophysiological models of depression suggest that overactivation of this amygdala-centered network underlies the negative attention and memory biases in depression (94). We are also particularly interested in studying habituation of the amygdala response which has been shown to correlate negatively with anxiety (95) and which is decreased in autism spectrum disorder (96-98). Both mean amygdala activation and habituation have been frequently used in genetic imaging studies to investigate the neural effects of genetic variants that are linked to depression, anxiety and personality traits like neuroticism (98, 99). For example, the short allele of the serotonin transporter gen has been associated with increased risk for depression after exposure to stress, which is thought to be mediated by increased amygdala reactivity to threat (99). Another line of evidence indicates that early life adversity is related to enhanced corticolimbic reactivity to negative facial expressions which is in turn related to rumination, which is known to be a vulnerability factor for internalizing psychiatric disorders (100).

### Data handling

We will store raw and cleaned data in a digital research environment (DRE). Data is also shared with researchers via the DRE. A variety of analysis software and statistical programs will be used to analyze the data. Statistical analysis will be performed within e.g. SPSS (version 25) and R (Project for Statistical Computing; version 3.6.1). Analysis of neuroimaging data will be performed with e.g. FSL (FMRIB Software Library version 5.0) for connectivity analyses before and after stress induction, SPM12 (Statistical Parametric Mapping version 12) for the emotional face matching task and Freesurfer (version 6.0.0) for analysis of the structural MRI and diffusion data. Data will be analyzed according to the state-of-art analyses insights and using relevant new techniques and approaches where applicable.

Digitalized diagnostic interviews are used in order to facilitate completeness of the diagnostic data. A data manager coordinates the data entry in the digital research environment, while also checking data quality. Data archiving and creating variables and scales is part of data management. Yearly study monitoring is carried out by an independent monitor to assess adherence to the procedures and to ensure patient safety and privacy.

### Statistical analysis plan

#### Distributions and missing data

In general, non-normally distributed data will be transformed, if possible. If the distribution remains non-normal after transformation, non-parametric tests will be applied. Missing data will be handled by multiple imputations, in the case of at random missing data and exceeding >5% missing data.

### Behavioral and physiological data analysis

All behavioral data from neuropsychological tests and self-report data from questionnaires will be scored according to published guidelines. “Classical” analyses will be carried out using general linear models such as AN(C)OVAs for comparisons between groups and regression, correlational and mediation analyses. These data will also be integrated into the analysis of neuroimaging and biological data, e.g. by means of regressors for first-level analyses within the framework of the general linear model (GLM), a behavioral regressor of interest in a higher level (cross-subject/group) comparison, or network analyses. In addition, we will employ data-driven multivariate approaches to discern latent variables across the different units. Principle component analyses (PCA) will be carried out at each unit of analysis to extract components from the self-report, behavior and physiological data, to be used in canonical correlation analyses.

### Resting state fMRI analysis

Resting state analyses are performed on the three resting-state scans at 1) baseline, 2) after the neutral and 3) after the aversive movie using FSL 5 (101).

#### Preprocessing

The scans are preprocessed using the FMRI Expert Analysis Tool (FEAT), which is part of the FMRIB Software Library (FSL). To account for T2* equilibration effects, the first five images of each resting-state scan are discarded. Further preprocessing steps include brain extraction, motion correction, bias field correction, high-pass temporal filtering with a cut-off of 100 seconds, spatial smoothing with a 4 mm full width at half maximum (FWHM) Gaussian kernel, registration of functional images to high-resolution T1 using boundary-based registration and nonlinear registration to standard space (MNI152). The final voxel size for group analysis is 2 mm isotropic. ICA-based Advanced Removal of Motion Artefacts (ICA-AROMA) is used for further single-subject denoising (102). Subjects are excluded from analyses if motion results in more than 2 mm sudden relative mean displacement or translation.

#### First level analyses (subject level)

Group (temporal concatenation) independent component analysis (ICA), implemented in MELODIC (103), is used to decompose the data of all subjects together into 20 components.. Network templates of the networks of interest (DMN and CEN (104), SN (105)) are used to select the relevant network components from the group ICA in the decomposed data of the first baseline resting-state scan across all subjects. For the whole brain analysis we generate subject-wise statistical maps of each of the networks of interest using dual regression. Regression of these time courses against the data resulted in spatial maps of the four networks of interest for each individual subject (106). Moreover, individual, aggregate measures of within network connectivity are generated by extracting the network connectivity during resting-state scan 2 and 3 from thresholded statistical masks of our networks (Z ≥ 3). The changes in network connectivity are investigated in parallel to the changes in our behavioral measures (subjective stress, heart rate and cortisol)

Next to this more static approach to connectivity, we will employ dynamic connectivity analyses, based on a sliding window approach and k-means clustering to derive functional connectivity states (106). Moreover, dynamic measures of network cohesion are derived to allow for real-time analyses in which physiological indices during stress induction can be integrated (107). The basic idea is that dynamic functional connectivity data along the baseline-stress-recovery axis for the three distinct networks will translate into functional differences between patients, with distinct symptom profiles and patterns of neuropsychological functioning.

### Structural MRI analysis

T1 images are analysed with FreeSurfer 6.0 (108) which provides a full processing stream for structural MRI data, including skull stripping, B1 bias field correction and gray-white matter segmentation. It results in a reconstruction of cortical surface models (i.e. cortical thickness) as well as a segmentation of subcortical brain structures (i.e. volume of hippocampus and amygdala).

Diffusion-tensor imaging data are analysed with TRACULA (TRActs Constrained by UnderLying Anatomy), a tool implemented in Freesurfer 6.0 for the automatic reconstruction of white-matter pathways, using global probabilistic tractography with anatomical priors. Prior distributions on the neighboring anatomical structures of each pathway are derived from an atlas and combined with the FreeSurfer cortical parcellation and subcortical segmentation of the subject that is being analyzed to constrain the tractography solutions (109).

It also generates voxel-wise estimates of fractional anisotropy or mean diffusivity, that, along with the volumetric and cortical thickness can be compared within and across groups/conditions by means of parametric or non-parametric tests (e.g. t-tests of effect size difference) within the framework of the GLM.

### Cross-sectional analyses

Our ultimate goal is to relate features of the different units of analysis across the different domains with multivariate methods. Extracted components from the self-report, behavior and physiological data are used as inputs in regularized canonical correlation analyses, to detect connections among the different units of analysis and identify transdiagnostic patterns in the data.

In addition, we will adopt a normative modeling approach for mapping associations between brain function, biological and clinical measures, and behavior and to estimate deviation from the normative model on a subject level. Normative modelling provides a framework to characterize patients individually in relation to normal functioning, which may be far more informative than categorical labels. This approach may help to parse the heterogeneity that is common in clinical cohorts and point to more biologically valid subtypes (110).

#### Microbiome data

We will apply a multi-step approach aimed to properly identify bacterial taxa, determine their co-occurrence patterns and assess its association with MRI-derived traits, behavior and cognition. First, we will perform a filtering procedure in order to remove low abundance genera and to reduce the impact of the high number of absent **o**perational taxonomic units (OTUs)/genera (with zero value per sample) where it is not possible to disentangle if these are the result of true zeros. Second, we estimate within-sample diversity metrics (four alpha-diversity metrics) and between-sample diversity metrics (β-diversity). Third, composition analysis and regression of taxonomic data (from phylum to genus) by 1) identifying which bacterial groups are present in our sample and their relative abundance, 2) establishing their genetic structures and 3) applying phylogenetic methods to establish their biological relationship. Fourth, we want to identify the underlying (functional) networks present in the microbiome using community structure detection algorithms. Fifth, characterize key interactions within the microbiome. Sixth, test and identify gene ×environment interactions between the microbiome and relevant genetic signaling systems.

### Dissemination

The study results will be published in peer-reviewed journals and distributed via media outlets. We will post our preprints at bioRxiv, a free online archive and distribution service for unpublished preprints in the life sciences. It is operated by Cold Spring Harbor Laboratory, a not-for-profit research and educational institution. By posting preprints on bioRxiv, Mind Set authors are able to make their findings immediately available to the scientific community and receive feedback on draft manuscripts before they are submitted to journals. Results will further be presented at national and international congresses and meetings. Participants are notified of study progress and outcome by means of newsletters.

## Data Availability

The datasets generated during and/or analyzed during the current study are not publicly available due to privacy reasons but are made available for researchers within the digital research environment (DRE) upon reasonable request to the corresponding author and approval of the steering board of the MIND-SET study group.

## Declarations

### Ethics approval and consent to participate

#### Regulation statements

The MIND-Set study has been approved by the local medical ethical committee (‘Commissie Mensgebonden Onderzoek Arnhem-Nijmegen’). After verbal and written information about the study which they received at home, eligible participants are approached by their care provider for participation in the study. If interested, they sign an informed consent form. A trained research professional schedules the test sessions with the participants. In the information it is clearly indicated which information would be gathered for clinical proposes and which measures were only part of the MIND-Set study. Written informed consent is provided for clinical data use and data collection. In the course of the study a yearly data monitoring is conducted with a local monitor of the Radboudumc Nijmegen

All diagnostic interviews, neuropsychological measures, physiological measures and neuroimaging measures are conducted by extensively trained clinicians and research assistants. All clinicians received diagnostic interview training from certified and experienced trainers. All research professionals conducting the neuropsychological tests received an extensive training by neuropsychological testing experts.

#### Compensation

Participants were compensated with travel costs for the data collection sessions and the controls were as well as were paid a small fee for their participation according to the guidelines of the medical ethical committee.

### Consent for publication

Not applicable

### Authors’ contributions

PE RC JV AA IT and AS conceived the design of the study and the study protocol, PE RC DG JV IT and AS wrote the first draft of the manuscript. PE RC JV AA EM SB FD MB JO IT AS revised the manuscript. All authors approved the final manuscript

## Acknowledgements

Not applicable

## Footnotes

1 During the preparation of this study the transition from DSM-IV to DSM-5 was still ongoing. Dutch translations of questionnaires and interviews according to DSM-5 were only limitedly available and if so psychometric testing was not yet completed, therefore the DSM-5 criteria could only be used for the diagnosis of ASD.

