## supplemental material for "Measuring Integrated Novel Dimensions in Neurodevelopmental and Stress-related Mental Disorders (MIND-Set): a cross-sectional comorbidity study from an RDoC perspective"

**SUPPLEMENTARY MATERIAL**

**PRE-ASSESSMENT**

**Demographics standard questionnaire**

We measure sex, age, ethnicity, religion, native language, age of parents at birth, marital status, living arrangements, level of education, work and religion with a questionnaire from the Dutch Helius study (1).

**Family Interview for Genetic Studies (FIGS)**

For the genetic studies, we use an instrument to categorize the family history of psychiatric disorders. The FIGS is a guideline originally used to gather information on the relatives of patients with schizophrenia and bipolar disorders and was developed by Gershon et al. (1988) and Maxwell (1992) (2, 3). Subjects are asked to provide diagnostic information about relatives in the pedigrees being studied (3).

**Adult ADHD Self-Report Scale Screener (ASRS)**

The screening version of the Adult ADHD Self-Report Scale (ASRS) Version 1.1 is a 6-question scale with good psychometric properties designed to screen for adult ADHD in community samples (4). The scale is short, easily scored, and can detect the vast majority of adult attention-deficit/hyperactivity disorder cases in the general population with high sensitivity and specificity, discriminating well among patients presenting for evaluation and specialty treatment (5). The cut-off for positive screening of ADHD is >3. Only when a patient scores above this cut-off, an additional diagnostic interview will be conducted during the clinical examination.
 **Conners’ Adult ADHD Rating Scale (CAARS)**
The CAARS intends to measure current ADHD symptomatology. It was developed for standardized self-ratings from adults undergoing evaluation for ADHD (6). Even though questions are being raised about its internal consistency, especially for the Hyperactive/Impulsive Symptoms subscale when completed by women, the CAARS was chosen as it is currently the most used test to measure adult ADHD symptoms for research and clinical purposes (7). **Autism-spectrum Quotient (AQ-50)**

The AQ-50 is a widely used 50-item self-report questionnaire, which was originally developed as a screening tool for ASD in psychiatric clinical populations and in the general population (8-10). It measures the degree to which an individual of normal intelligence shows “autistic traits”, or traits associated with the autistic spectrum. The Dutch translation was validated by Hoekstra et al. (9).

The AQ-50 can be used for screening purposes and as a measure of symptom severity in ASD. Only when a patient scores above the cut-off (>25) of the AQ-50 for autism spectrum disorders, an additional diagnostic interview will be conducted during the clinical examination.

**Inventory of Depressive Symptomatology self-report version (IDS-SR)**

Severity of depressive symptoms is measured with the 30-item IDS-SR, and comprises three factors: cognition and mood, anxiety and arousal, and sleep and appetite regulation. Items are scored on a four-point scale (range 0-3), with each item equally weighed. 28 of the 30 items are summed to a standard total score, ranging from 0 to 84 (higher score = more severe depression). Scores ≤13 are considered within the normal range; scores of 14-21 indicate mild MDD, 22-38 moderate MDD and ≥39 severe MDD. The IDS-SR has highly acceptable psychometric properties (11).

**Anxiety Sensitivity Index (ASI)**
As a measure for anxiety we use one of the most widely used instruments to quantify anxiety sensitivity (AS), which refers to the fear of anxiety-related physical sensations because of beliefs that these sensations may have potentially harmful physical, social or cognitive consequences (12, 13). As a trait variable, AS is considered an underlying cognitive construct that has been linked to cognitive and behavioral theories as well as theoretical explanations for the development of all anxiety disorders as well as depression. The 16 item ASI shows good psychometric properties (14) and contains a three factor structure: physical concerns, mental incapacitation concerns and social concerns (15).

**Personality Inventory for DSM-5 - Brief Form (PID-5-BF)**- Adult

We use the Dutch version of PID-5-BFto measure the trait part of the alternative model for personality disorders proposed in Section III of the DSM-5 (16). This PID-5-BF-NL is a 25-item self-rated personality trait assessment scale for adults aged 18 and older (17). It assesses five personality trait domains including negative affect, detachment, antagonism, disinhibition, and psychoticism, with each trait domain consisting of 5 items. Each item is rated on a 4-point scale (i.e., 0=very false or often false; 1=sometimes or somewhat false; 2=sometimes or somewhat true; 3=very true or often true). The overall measure has a range of scores from 0 to 75, with higher scores indicating more severe overall personality dysfunction. Each trait domain score ranges from 0 to 15, with higher scores indicating greater dysfunction in the specific personality trait domain. Besides the total score, average scores can be calculated for each domain and for the overall measure. The average scores reduce the overall score as well as the scores for each domain to a 4-point scale, which allows the clinician to assess the patient’s personality dysfunction relative to observed norms. The average domain score is calculated by dividing the raw domain score by the number of items in the domain (e.g., if all the items within the “negative affect” domain are rated as being “sometimes or somewhat true” then the average domain score would be 10/5 = 2, indicating moderate negative affect). The average total score is calculated by dividing the raw overall score by the total number of items in the measure (i.e., 25). The average domain and overall personality dysfunction scores were found to be reliable, easy to use, and clinically useful to the clinicians in the DSM-5 Field Trials (18).

**General health: Short-Form-20 (SF-20)**

General health is measured with the Short-Form-20 (SF-20). The SF-20 is a multipurpose, short-form health survey that contains 20 questions (19). The SF-20 has three dimensions both for functioning (physical, social and role) and for well-being (mental health, health perception and pain). It yields six numerical scores for each parameter such that a higher score indicates better functioning or wellbeing. The only exception is pain, a higher score indicates more pain. The SF-20 has been validated in adult and elderly populations (20, 21), and shows adequate reliability and validity (22).

**WHO-Disability Assessment Schedule 2.0 (WHODAS)**

Functional limitations are assessed with the WHODAS 2.0, which consists of 36 questions, covering six domains of functioning during the last 30 days: 1. Cognition (understanding and communicating); 2. Mobility (moving and getting around); 3. Hygiene, dressing, eating and staying alone (self-care); 4. Interpersonal actions (getting along with people); 5. Work, leisure, domestic responsibilities (household activities); 6. Joining in community activities (participation in society) (23). This self-report questionnaire integrates an individual's level of functioning in major life domains and directly corresponds with the ICF "activity and participation" dimensions. After calculating the total scores and total domain scores, the scores were transformed into adjusted scores on a scale from 0 to 100, where higher scores reflect greater disability (24).  **Health related** **quality of life: Outcome Questionnaire (OQ-45.2)**
We measure quality of life with the Dutch version of the Outcome Questionnaire (OQ-45.2), a validated scale with good psychometric properties (25, 26). The scale consists of 45 items scored on a scale from 0 to 5 reflecting current (dys)functioning. Along with a total score ranging from 0 to 180, three subscales can be calculated: symptom distress, interpersonal relationships and social role, with higher scores indicating more problems.

**SESSION 1: CLINICAL EXAMINATION**

A. **Psychopathology diagnostics**

Trained clinicians will conduct the diagnostic interviews and experienced psychiatrists will supervise the diagnostic procedure closely. Difficult assessments are discussed and in the case of disagreement we will make a conservative decision (e.g. exclusion).

The study takes place during the DSM-IV/-5 transition period. For classification according to DSM-IV or DSM-5 we use the following structured interviews:

***Neurodevelopmental disorders***

Neurodevelopmental disorders are assessed during a two-step diagnostic screening procedure, using the ASRS for ADHD screening and the AQ-50 (see above) for ASD screening. When screening questionnaires are scored positive, the NIDA will be administered in order to diagnose ASD and/or the DIVA will be administered in order to diagnose ADHD. Both the DIVA and NIDA are completed in the presence of a partner and/or family member of the patient to be able to retrospectively and collaterally ascertain information on a broad range of symptoms in childhood and adulthood.

**Diagnostic Interview for Adult ADHD, second edition (DIVA 2.0)**

To diagnose or classify ADHD we use the DIVA, a semi-structured interview that can be used to classify according to both DSM-IV and DSM-5 criteria and can be conducted by trained clinicians (27).

**Dutch Interview for autism spectrum disorders in adults (NIDA)**

The NIDA (28) is an interview to classify and diagnose autism spectrum disorders in adults according to DSM-5 criteria. The NIDA is a semi-structured interview that can be administered by trained clinicians.

***Stress-related disorders***

**Structured Clinical Interview for DSM-IV Axis I Disorders (SCID-I)**

To diagnose mood disorders and anxiety disorders according to DSM-IV we use the appropriate sections of the SCID-I, a clinical interview designed for use in clinical and research settings, with a high validity for depressive and anxiety disorders (29). The psychotic disorders section is also conducted in order to exclude patients with a current psychosis from participation.

**Measurements in the Addictions for Triage and Evaluation and criminality (MATE-Crimi)**

To diagnose/classify substance related disorders according to DSM-IV and DSM-5 we use the Addictions for Triage and Evaluation and criminality (MATE-crimi; subsections: 1, 3, 4, 9 and Q1). The MATE assesses the use of psychoactive substances and the level of craving (30).

B. **Health markers**

**Presence of somatic diseases**

Somatic co-morbidity is assessed by the Health Interview Questionnaire (31): a self-report questionnaire about the presence of somatic diseases, including: Lung disease (asthma, chronic bronchitis, pulmonary emphysema), cardiovascular disease (cardiac events, heart failure, heart infarction, cardiac arrhythmia, coronary heart disease, angina pectoris, vascular abnormalities), stroke, diabetes, arthritis/ rheumatism (osteoarthritis, rheumatism), gastro intestinal disease (ulcer, irritable bowel syndrome, Crohn’s disease, colitis ulcerosa, constipation), cancer, epilepsy, thyroid disease, or any other disease. Compared to general practitioner information, the accuracy of the questionnaire was shown to be adequate and independent of cognitive impairment (32).

**Medication use**

Current medication use is determined by medication verification from at least 2 sources: The list of medication that the patient brings from the pharmacy or container inspection was observed and combined with questions about current use.

**Physical examination**

The physical examination, including height, weight, waist circumference, BMI, pulse rate and blood pressure measurements (the latter in both lying and standing position) and visual acuity, was conducted during the first clinical appointment.

C. **Biological markers**

**Blood sample**

These are taken during the clinical appointment. A trained nurse draws four additional venous blood samples (1 x 10 ml EDTA for DNA, 1 x 10 ml EDTA for plasma, 1 x 10 ml STOL for serum, and 1 x 2.5 ml PAX for RNA) , according to standard procedures. Blood is immediately transported at room temperature to a local laboratory. Within 72 hours after withdrawal, the blood sample will be registered by the facility, given a unique number code and will be stored at -80°C until further use. Assays included albumin, creatinine, gene expression (DNA, RNA, Epigenetics), glucose level, HDL & LDL cholesterol, haemoglobin, hematocrit, inflammatory markers, kidney function, liver function, thyroid function and triglycerides.

**Faeces microbiome**

Patients are asked to provide their stool sample following standard protocols. Patients collect the sample at home through a faeces collection kit and sent it to the department of Human Genetics at Radboudumc via regular mail in a secure envelope. Samples are stored at -80°C until sequenced. Bacterial DNA will be isolated and the 16S rRNA gene is sequenced. Taxonomic identification and abundances of different bacteria are determined or clustered into operational taxonomic units at a certain similarity level in a taxonomic independent way. At 97% similarity level, these OTUs are used to approximate the taxonomic rank species.

**Hair sample**

A hair sample (<5 mm in diameter, >3 cm length) will be taken from the participants and five additional questions about hair care are asked in order to be able to control for potential confounding factors. Hair cortisol values will subsequently be analyzed by enzyme immunoassay in order estimate long-term stress exposure (33).

**SESSION 2: BEHAVIOURAL SESSION**

A. **Questionnaires**

**NEMESIS-childhood trauma questionnaire**

This structured inventory, developed for the NEMESIS epidemiological study in the Netherlands, retrospectively measures an individual’s trauma history before the age of 16 (34, 35). The questionnaire allows for the identification of four meaningful types of traumatic childhood events within the family: emotional neglect, psychological abuse, physical abuse, and sexual abuse. The frequency of each type of childhood trauma is scored on a five-point Likert scale, ranging from “0” (not) to “5” (very often). The “childhood trauma index” (36) is calculated by taking the sum of the frequency scores of the four types of traumatic events and represents a combination of frequency and diversity of traumatic events. This index is highly correlated with the severity of psychopathology is regarded as a strong variable for examining the association between childhood trauma and psychopathology (37).

**Food intake questionnaire**

A diet-lifestyle questionnaire originally developed for TACTICS (38) is assessed to report on patterns of food intake relevant for microbiome analyses.

**Toronto Alexithymia Scale-20 (TAS-20)**

The TAS-20 is an established self-report measure of alexithymia. The items are scored on a 5-point likert scale. The scale has three subscales: Difficulty describing feelings (5 items; range: 5-25), Difficulty identifying feelings (7 items; range: 7-35), and Externally-oriented thinking (8 items; range: 8-40). The total alexithymia score (range 20-100) is interpreted as non (<51), possible (51-60), or alexithymia (>60) (39).

**Behavior Rating Inventory of Executive Functioning –Adult version (BRIEF-A)**

We use an adapted version of the BRIEF-A, a valid measure to assess functional problems related to everyday executive functioning. The questionnaire consists of 75 standardized items, of which items measure different aspects of executive function within nine clinical scales (Inhibit, Shift, Emotional Control, Self-Monitor, Initiate, Working Memory, Plan/Organise, Organisation of Materials, and Task Monitor). The nine scales are separated into two indices: the Metacognitive Index (MI) which consists of five scales (Task Monitor; Organization of Materials; Plan/Organize; Working Memory and Initiate) and the Behavioural Regulation Index (BRI) which contains four scales (Emotional Control; Shift; Inhibit and Self-Monitor). The two indices sum up to one composite summary score (GEC). Internal consistency is high with alpha coefficients ranging from .80 to .98, and high test-retest reliability ranging from .91 to .94 (40).

**Perseverative Thinking Questionnaire (PTQ)**

The perseverative thinking questionnaire (PTQ) (41) is a 15-item self-report questionnaire used to estimate patients’ level of repetitive negative thinking from a transdiagnostic perspective. Items are rated on a 5-point scale ranging from 0 (never) to 4 (almost always). Studies confirm the adequate psychometric characteristics of the PTQ (41, 42). Three lower-order factors can be distinguished, namely 1) core characteristics of perseverative cognition (repetitiveness, intrusiveness, difficulties to disengage; e.g., “The same thoughts keep going through my mind again and again”), 2) perceived unproductiveness of perseverative cognition (e.g., “My thoughts are not much help to me”), and 3) mental capacity captured by perseverative cognition (e.g., “I can’t do anything else while thinking about my problems”).

**Relation self and problems**

The following single question is used to assess relation with self and mental problems: “To what extent are your mental problems a part of you?”

B. **Neuropsychological tests**

The neuropsychological tasks are presented on a computer and a tablet in a quiet dimly lit sound proofed room where dimming of the light was adapted to the need of the subjects and the tasks. There is one paper-and-pencil task. We measure cognitive bias (as part of emotional processing) using an attentional bias task, an emotion recognition task and two memory bias tasks. Several tests are included to measure different aspects of executive functioning, as well as a test to estimate premorbid intelligence. Visual analogue scales are used to assess mood at four different timepoints throughout the assessment, as well as self-reported effort on the tests afterwards.

**Self-referent memory bias**

After a first mood rating, the computerized Self-Referent Encoding Task (SRET) (43) is used to assess explicit negative memory bias. The SRET is a dependable test to measure memory bias and has frequently been used in other memory bias studies (37, 43-45). The SRET consists of an encoding phase and a recall phase. During the encoding phase, twelve positive and twelve negative possibly self-descriptive adjectives are individually presented on a computer screen in a fixed randomized order. These words were aimed at triggering positive and negative cognitive schemas (46). The valence of these words was confirmed by 99 independent volunteers (79% female, M age = 29 years, SD age = 15.12 years) who rated all words on a scale of 1 (extremely negative) to 10 (extremely positive). The valence of the positive words (M = 6.02, SD = 0.49) was significantly more positive than the valence of the negative words (M = 2.69, SD = 0.72), t(98) = -34.57, p < .001. Participants are instructed to indicate how well each word describes them on a five-point Likert scale ranging from one (not very well) to five (extremely well). After a two-minute distraction task (the Coding subtest from the WAIS-IV-NL) (47), a free recall phase follows. The SRET is followed by a second mood rating.

**Prepotent response inhibition, interference control and alertness**

Several subtests from the normed Testbatterie zur Aufmerksamkeitsprüfung Version 2.3 (TAP 2.3) are administered. The TAP 2.3 is a widely used, reliable computerized test battery whose validity has been established in a range of clinical populations (48). The Go/Nogo subtest is used as a measure of prepotent response inhibition, the Incompatibility subtest to measure interference control and incompatibility (the “Simon effect”), and the Alertness subtest to assess intrinsic and phasic alertness.

**Intelligence**

Premorbid IQ is estimated using the Dutch version of the National Adult Reading test, the “Nederlandse Leestest voor Volwassenen”(NLV) (49). **Reversal learning**

We employ a probabilistic reversal learning task (50-52) to examine reward sensitivity. In this task participants have to choose one out of two available squares, after which they receive feedback – either a reward or punishment – on their choice. This feedback is given in a probabilistic fashion, 80% of the feedback is correct. After a while the contingencies change; the square that was previously rewarded is now punished, and vice-versa. With this task it is possible to measure on a trial-by-trial basis whether people repeat behavior that was previously rewarded. This is called the win-stay rate, which is an index of reward sensitivity. A higher win-stay rate indicates increased reward sensitivity, as participants choose the same option more often after receiving a reward. This measure allows us to examine possible differences in reward sensitivity between different disorders.

**Spatial working memory** **and shifting**

Three subtests from the normed Cambridge Neuropsychological Test Automated Battery (CANTAB) (53) are administered on a tablet. This test battery has a particularly strong theoretical basis and its validity has been tested in a wide range of neuropsychological and psychiatric patients (54). The subtest Spatial Working Memory is used to measure spatial working memory and the Intra-Extra Dimensional Set Shift subtest is used to measure rule acquisition and reversal. The Motor Screening Task is included to check the validity of assessment using the tablet (53).

**Attention bias**

A non-invasive computer-mounted ‘beam’ eye-tracking system (55) is used for the following attentional bias task. The accuracy of the eye tracker is 0.4◦, with a processing rate of 500 Hz. The direction of gaze is measured in x and y coordinates. Pictures were selected to fit three categories, presented in different blocks in randomized order (4 pictures per trial presented in a grid): 1. social content (i.e., one person or more individuals), 2. non-social content (i.e., animals and landscapes), 3. facial expressions (happy, neutral, sad, angry). For both the social and non-social block, half of all the stimuli are positively and the other half negatively valenced. Each trial begins with a 1000ms centrally presented fixation cross, followed by the simultaneous presentation of four pictures for 30s. A total of 12 trials per block are presented. Participants are instructed to freely view the pictures on the screen. To stimulate active viewing, participants are told that a memory test will follow.

**Memory bias for eye-tracking task stimuli**

During the eye-tracking memory test, two out of four pictures from the grids presented during the attention bias task have switched position and participants are asked to indicate one of the relocated pictures by clicking on it using a mouse. Attention focus is measured throughout.

**Emotion recognition**

While attention focus is assessed using the eye-tracking system, recognition of facial expressions is assessed. New (i.e. not in the attention bias task) pictures of individuals presenting different facial expressions of emotional expressions are presented centrally on the screen for 30s. The stimuli included a total of 96 pictures selected from the International Affective Picture System (IAPS) and the Nencki Affective Picture System (NAPS) (56). Participants are instructed to indicate which emotion (happy, neutral, sad, angry) best fits the facial expression as accurately as possible using corresponding keyboard keys.

**Self-rated mood and motivation**

At four different timepoints during the session, mood is assessed using four Visual Analogue Scales (“How negative, sad or bad are you feeling at this moment?”; “How positive, happy or good are you feeling at this moment?”; “How tense, agitated or restless are you feeling at this moment?”; “How relaxed or calm are you feeling at this moment?”). After all neuropsychological tests have been completed, motivation is similarly assessed using four Visual Analogue Scales (“I thought it was easy”; “I thought it was fun to do”; “I felt well rested”; “I did my absolute best”).

**SESSION 3: NEUROIMAGING SESSION**

All MRI scans take place between 12:00 and 22:00 in order to ensure low and relatively stable levels of endogenous cortisol. On the day of scanning, during a 45-minute pre-scanning acclimatization period, participants receive information about the study, fill in questionnaires and watched a relaxing nature documentary (57).

After locator and reference scans, a structural T1-scan will provide high resolution three-dimensional anatomical information. Then we will obtain a functional MRI scan during an emotional face matching task (58) to investigate amygdala reactivity and a resting-state scan to investigate connectivity patterns in large scale networks in the brain. After this first resting state scan, participants will watch a neutral movie clip from the movie 'Comment j'ai tué mon père' by Anne Fontaine (59), followed by a resting state scan. This is the control condition for the stress induction with a mild psychological stressor that follows. It consists of watching an aversive movie clip from the movie 'Irréversible' (2002) by Gaspar Noé (60), showing extreme male-to-male aggression and violence in front of a crowd. Participants were asked to constantly and attentively view the movie clips after a short introductory text put them in the scene from an eye-witness perspective, thereby involving them maximally in the experience. This paradigm closely corresponds to the determinants of the human stress response, i.e. unpredictability, novelty and uncontrollability (61). During acquisition of the resting-state data subjects are instructed to lie still with their eyes open and to keep their gaze fixed on a fixation cross while surrounding lights were dimmed. Participants are instructed not to think about anything in particular. This method has been shown to have the highest reliability and consistency in acquiring resting-state data (62) and promotes the subjects to remain awake. An MRI-compatible eye-tracking device using infrared light from SensoMotoric instruments (MEyeTrack-LR) is used check whether the participants indeed kept their eyes open. In combination with the resting-state scan after the first neutral movie clip, this will allow assessment of stress-induced changes in brain network interactions. Finally, a diffusion-weighted imaging (DTI) spin echo sequence will assess white matter structure.

All images were collected using a 3T Siemens Magnetom Prisma MRI scanner (63) with a 32-channel head coil. Six saliva samples were collected with a salivette on the scanning day, in order to measure the cortisol levels and get an indication of the stress level. Participants were also asked to collect two saliva samples at home, which were used to determine the baseline cortisol levels.

**Structural scan**

High-resolution structural images (1 mm isotropic) are acquired using a T1-weighted MP-RAGE sequence (TE/TR = 3.03/2300 ms, flip angle = 8°, FOV = 256xó256x192 mm, GRAPPA acceleration factor 2). Acquisition time ~5 minutes.

**Amygdala Reactivity Paradigm**

During scanning, participants performed an emotional face matching task consisting of a blocked design, including an emotion condition, and a visuo-motor control condition. This paradigm has been used previously to investigate drug effects on amygdala reactivity (58, 64). It robustly engages an amygdala-centered network by contrasting the response to simultaneously presented angry and fearful face stimuli with the response to geometric shapes (i.e. ellipses that consisted of scrambles of the same face stimuli). Two emotion blocks were interleaved with three control blocks, and each 30 s block consisted of six 5 s trials. Each trial consisted of three simultaneously presented stimuli, with the cue stimulus presented above the target and distractor. In the emotion condition, an angry or fearful face was presented on top as cue, and subjects had to indicate by an appropriate button press, which of the bottom two faces (one angry and one fearful) matched the cue in emotional expression. The three simultaneously presented faces per trial were from different persons from the same sex. Half the trials presented faces of men and half of women, half of each target emotion (angry or fearful). In the sensorimotor control condition, a horizontally- or vertically-oriented ellipse was presented as cue above two ellipses (one vertical and one horizontal), and subjects had to select the identically oriented ellipse. Note that by contrasting neural responses to mixed, negatively valenced face stimuli to geometric shapes, this task does not probe any emotion-specific effects, but rather the reactivity of (para)limbic affective circuitry to biologically salient, environmentally relevant stimuli.

**Resting state fMRI**:

Rs-fMRI is acquired before (baseline rs fMRI; 500 volumes), and after the neutral movie clip (500 volumes) and after the aversive movie clip (750 volumes). T2*-weighted EPI BOLD-fMRI images are acquired for the resting-state scans, using a multi-band 6 protocol with an interleaved slice acquisition sequence (number of slices = 66, TR = 1000 ms, TE = 34 ms, flip angle = 60¬∞, voxel size = 2.0√ó2.0√ó2.0 mm, slice gap = 0 mm, FOV = 210 mm).

**DTI**

High angular resolution diffusion imaging (HARDI) with 61 diffusion directions will be obtained (Field Of View (FOV): 200 x 257 x 126 mm, 60 slices, no gap, spatial resolution: 1.8 x 1.8 x 2.1 mm, TR / TE = 12561 / 59 ms, flip angle = 90°, half k-space acquisition will be used (half scan factor = 0.68), SENSE parallel imaging factor = 2.5, b-values = 0, 1200 s/mm2, with SPIR fat suppression and dynamic stabilisation in an image acquisition time of 15 min 42 s).

16.	American Psychiatric Association. Diagnostic and statistical manual of mental disorders (5th ed.). Washington, DC: American Psychiatric Association; 2013.

17.	Van der Heijden P, Ingenhoven T, Berghuis H, Rossi G. Nederlandstalige bewerking van The Personality Inventory for DSM-5 ® — Brief Form (PID-5-BF) — Adult, 2011. Amsterdam: Uitgeverij Boom; 2014.

25.	de Jong K, Nugter MA, Lambert MJ, Burlingame GM. Handleiding voor afname en scoring van de Outcome Questionaire OQ-45.2. Heiloo: GGZ Noord-Holland-Noord; 2008.

26.	Lambert MJ, Morton JJ, Hartfield D, Harmon C, Hamilton S, Reid RC, et al. Administration and Scoring Manual for the OQ-45.2 Outcome Questionnaire. Salt Lake City: American Professional Credentialing Services; 2004.

27.	Kooij J, Francken M. DIVA 2.0. Diagnostic Interview for ADHD in adults (DIVA). The Hague: DIVA Foundation; 2010.

28.	Vuijk R. Nederlands interview ten behoeve van diagnostiek autismespectrumstoornis bij volwassenen (NIDA). Rotterdam: Sarr Expertisecentrum Autisme/Dare to Design; 2016.

29.	First M, Gibbon M, Spitzer R, Williams J, Benjamin L. Structured clinical interview for DSM-IV-TR axis I disorders, research version, patient edition. (SCID-I/P). New York: Biometrics Research, New York State Psychiatric Institute; 1997.

30.	Schippers GM, Broekman TG, Buchholz A, Koeter MW, van den Brink W. Measurements in the Addictions for Triage and Evaluation (MATE): an instrument based on the World Health Organization family of international classifications. Addiction. 2010;105(5):862-71.

31.	Central Bureau of Statistics. Health Interview Questionnaire. Heerlen: CBS; 1989.

38.	TACTICS. Translational Adolescent and Childhood Therapeutic Interventions in Compulsive Syndromes [Available from: <https://www.tactics-project.eu/>.

46.	Beck AT, Rush AJ, Shaw BF, Emery G. Cognitive Therapy of Depression. : Guilford Press; 1987.

47.	Wechsler D. WAIS-IV-NL Afname- en Scoringshandleiding. 1 ed. Enschede: Ipskamp Drukkers B.V.; 2012.

48.	Zimmerman P, Fimm B. TAP 2.3 - Testbattery for attentional performance. Herzogenrath: Psytest; 2012.

49.	Schmand B, Lindeboom J, van Harskamp F. NLV: Nederlandse leestest voor volwassenen. Lisse: Swets & Zeitlinger; 1992.

55.	SMI RED500: iMotions; [Available from: <https://imotions.com/hardware/smi-red500/>.

56.	Marchewka A, Żurawski Ł, Jednoróg K, Grabowska A. The Nencki Affective Picture System (NAPS): Introduction to a novel, standardized, wide-range, high-quality, realistic picture database. Behavior Research Methods. 2014;46(2):596-610.

57.	Attenborough D. Life. 2009.

58.	Hariri AR, Mattay VS, Tessitore A, Fera F, Smith WG, Weinberger DR. Dextroamphetamine modulates the response of the human amygdala. Neuropsychopharmacology. 2002;27(6):1036-40.

59.	Fontaine A. Comment j’ai tué mon père. 2001.

60.	Noé G. Irréversible. 2002.

61.	Mason JW. A review of psychoendocrine research on the pituitary-adrenal cortical system. Psychosomatic Medicine. 1968;30(5, Pt. 2):576-607.

62.	Patriat R, Molloy EK, Meier TB, Kirk GR, Nair VA, Meyerand ME, et al. The effect of resting condition on resting-state fMRI reliability and consistency: a comparison between resting with eyes open, closed, and fixated. NeuroImage. 2013;78:463-73.

63.	MAGNETOM Prisma [Available from: <https://www.siemens-healthineers.com/nl/magnetic-resonance-imaging/3t-mri-scanner/magnetom-prisma>.

64.	van Wingen GA, van Broekhoven F, Verkes RJ, Petersson KM, Bäckström T, Buitelaar JK, et al. Progesterone selectively increases amygdala reactivity in women. Molecular Psychiatry. 2008;13(3):325-33.
